# Estimating time-dependent infectious contact: a multi-strain epidemiological model of SARS-CoV-2 on the island of Ireland

**DOI:** 10.1101/2022.03.25.22272942

**Authors:** Tsukushi Kamiya, Alberto Alvarez-Iglesias, John Ferguson, Shane Murphy, Mircea T. Sofonea, Nicola Fitz-Simon

## Abstract

Mathematical modelling plays a key role in understanding and predicting the epidemiological dynamics of infectious diseases. We construct a flexible discrete-time model that incorporates multiple viral strains with different transmissibilities to estimate the changing infectious contact that generates new infections. Using a Bayesian approach, we fit the model to longitudinal data on hospitalisation with COVID-19 from the Republic of Ireland and Northern Ireland during the first year of the pandemic. We describe the estimated change in infectious contact in the context of government-mandated non-pharmaceutical interventions in the two jurisdictions on the island of Ireland. We take advantage of the fitted model to conduct counterfactual analyses exploring the impact of lockdown timing and introducing a novel, more transmissible variant. We found substantial differences in infectious contact between the two jurisdictions during periods of varied restriction easing and December holidays. Our counterfactual analyses reveal that implementing lockdowns earlier would have decreased subsequent hospitalisation substantially in most, but not all cases, and that an introduction of a more transmissible variant - without necessarily being more severe - can cause a large impact on the health care burden.

## 1 Introduction

During an epidemic, behavioural changes are encouraged, and sometimes mandated, to curtail infectious disease transmission. These changes aim to reduce the number of contacts between people indiscriminately (e.g., closure of schools, workplaces, commercial establishments, roads, and public transit; restriction of movement; cancellation of public events; maintenance of physical distances in public) and reduce the chance of infection upon contact (e.g., use of personal protective equipment). Furthermore, tracing and isolating known infectious cases can limit the contact between infectious and susceptible individuals. These actions are collectively referred to as non-pharmaceutical interventions (NPIs) and are often mandated by governments. Slowing the surge of infection (or “flattening the curve”) affords an opportunity to reduce infection-induced mortality and mobility, alleviate health care burden and wait out an epidemic until pharmaceutical solutions (i.e., treatment and vaccines) become available. Implementation of mandated NPI in historical outbreaks, including during the 1918 influenza pandemic, was crucial for preventing excess death in the United States [1]. NPIs have also been mandated globally during the COVID-19 pandemic.

Mathematical modelling and quantitative analyses of empirical data plays a pivotal role in understanding and predicting epidemiological dynamics. Mechanistic epidemiological models have been widely applied to study the dynamics of SARS-CoV-2, and to make predictions of clinical outcomes under alternative scenarios (e.g., an assumed decrease in physical contact [2]). Despite their public health benefits, social distancing measures have been shown to incur high costs in several domains, including in economy [3], mental health [4], and civil liberty [5]. Thus, it is crucial to quantify infection contact, or its derivative quantities like the effective reproductive number, *R* to monitor changes in infection burden, achieve desired public health outcomes and improve policy transparency and public engagement. While it is not possible to measure infectious contact directly, fitting a mathematical model to longitudinal data on observed processes such as reported cases and hospital admissions allows estimation of inter-individual infectious contact and its derivatives [6,7].

A large contingency of epidemiological models follows a rich tradition of ordinary differential equation (ODE) models [8], which track the spread of infection and often immunity in a population. Specifically, ODE models assume that waiting time processes (such as infectious period and time to hospitalisation) are memoryless, that is to say, that the waiting time until an event (such as recovery and hospitalisation) does not depend on the elapsed time. Seen at the population level, this assumption deduces that times spent by individuals in each compartment are distributed exponentially, implying large individual variability. While mathematically convenient, the lack of memory is unsupported for certain epidemiological processes [9], and empirical evidence indicates other probability distributions with smaller individual variability and non-monotonic densities (e.g., gamma-, Weibull and log-normal distributions) are better equipped to describe those processes. Previous studies have also demonstrated that quantitative predictions of epidemiological outcomes depend on the assumed probability distribution in a variety of systems [10–13], including SARS-CoV-2 [7]. As such, it is pertinent to incorporate realistic waiting time distributions, particularly when one aims to obtain quantitative and short-term, rather than qualitative and long-term insights from epidemiological models.

Here we develop a compartmental epidemiological model that accurately predicts inter-individual infectious contact over the first year of the epidemic in the Republic of Ireland (ROI) and Northern Ireland (NI). These neighbouring jurisdictions on the island of Ireland present a compelling contrast due to independent policymaking over a small geographical area. We use a discrete-time approach, which allows us to incorporate more accurate assumptions about times spent in each compartment [7].

The COVID-19 pandemic has been characterised by subsequent waves of novel variants with varying disease transmissibility and severity. To separate the effect of human behaviour from the difference in transmissibility of multiple strains, we explicitly model multiple SARS-CoV-2 strains, with differing transmissibilities, seeded in the population at differing times. Our multiple strain model allows consistent estimates of relative infectious contact between periods even when the dominant variant has changed.

Previous studies of SARS-CoV-2 have estimated the time-dependent contact ratio and its derived quantities using continuous (e.g., basis splines [14]), or piece-wise, discrete functions of time (e.g., consisting of the specified period corresponding to NPI mandates [7]). However, both approaches can be fraught with challenges. On the one hand, it is not obvious to choose the appropriate extent of smoothing of a continuous function, for example, by deciding the number of knots in a basis spline function. On the other hand, abrupt changes imposed by piece-wise functions are at odds with empirical data on human movement during the COVID-19 pandemic [15]. Furthermore, it is difficult to establish a precise definition of the level of an intervention over time as definitions changed over time [16, HYPERLINK \l “bookmark23” 17]. To address these concerns, we develop an intermediate approach, in which we introduce a prior that allows smoothness in infectious contact between neighbouring weeks in the absence of information otherwise from empirical data.

In the Irish context, compartmental models have been used by several other research groups to understand the dynamics of the virus, make forecasts of outcomes under various scenarios, and assess economic impacts of policy restrictions [18–23]. Our study complements these studies by providing a high-resolution description of the change in infectious contact over time, comparing the two jurisdictions on the island of Ireland. Leveraging the epidemiological model and estimated parameters, we also perform counterfactual analyses to explore the effects of alternative interventions on cumulative hospitalisation and assess the impact of a novel variant.

## Methods

### Epidemiological model and data

Our multi-strain discrete-time model consists of three types of host compartments (Fig. 1): a susceptible compartment (*S*) and two infectious compartments for viral strain *s*, (*J*_*s,i*_ and *Y*_*s,i*_ where *i* indicates the infection age, i.e., day since exposure). The compartments *J* and *Y* differ in their future clinical outcome: individuals in the components *Y* eventually get hospitalised while those in *J* remain out of hospitals. As our primary focus is the inference of NPI in the community, we did not consider within-hospital transmission, recurring hospital admissions of the same patients, or demographic turnover. We ignored the dynamics of recovered hosts who were assumed to have minimal influence on the transmission during the period investigated.

**Figure 1:**
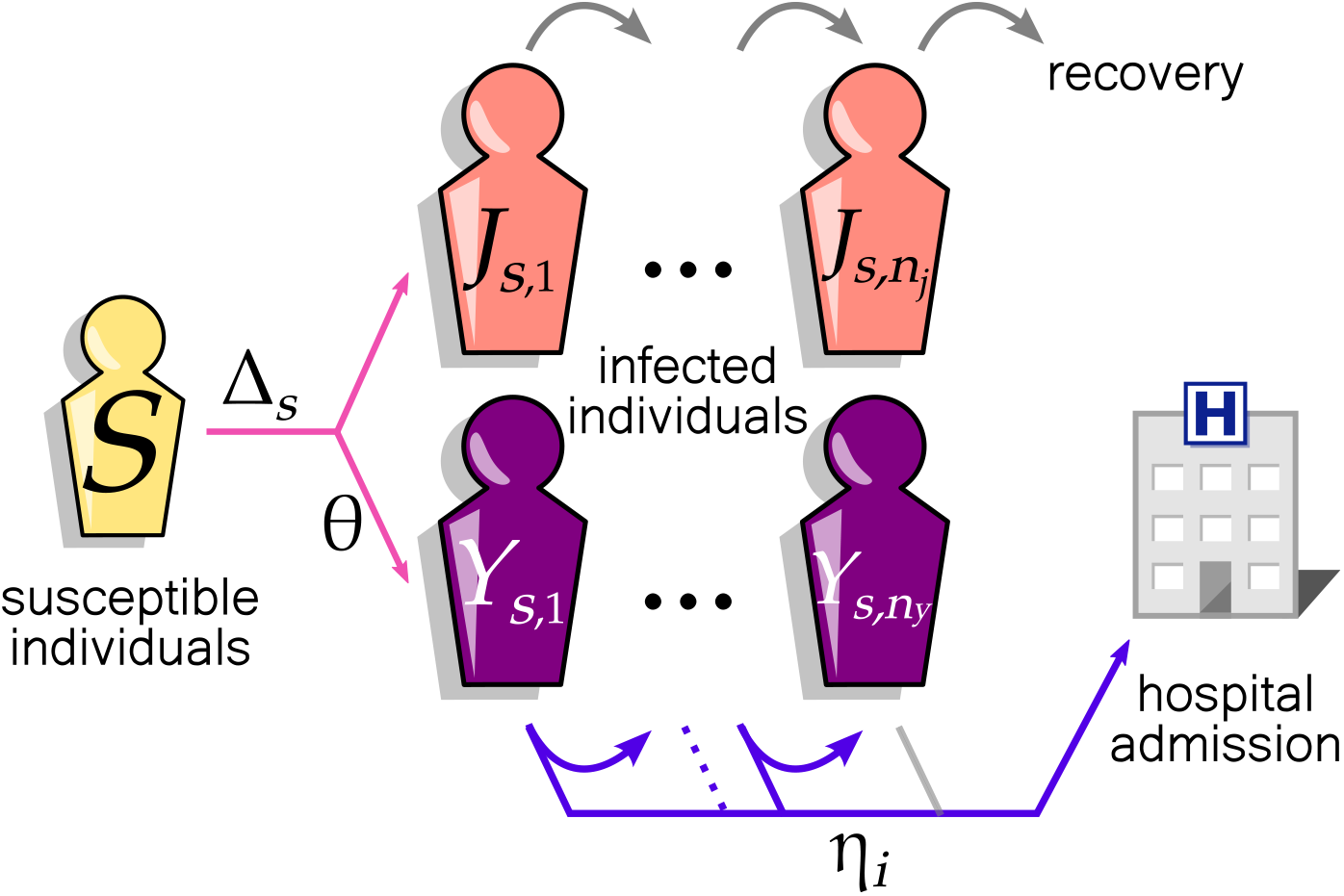
Discrete-time model of SARS-CoV-2 community transmission consists of susceptible *S* and infectious (*J* and *Y*) compartments of pathogen strain *s*. Each square represents a group of individuals with an identical contribution to the epidemiological dynamics. Infection with strain *s* occurs with probability Δ_*s*_ per day. Individuals in the components *Y* are infectious patients to be hospitalised. Once infected, individuals progress to the next square each day (*J* and *Y*), capturing the memory effect of the infection age. After spending *n*_*j*_ days, infectious hosts in *J* are no longer infectious. Alternatively, a fraction, *θ* of infectious hosts (in *Y*), is admitted to the hospital with a delay specified by the probabilities 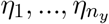, where *η*_*i*_ is the probability that the individual is admitted to hospital on the day *i*, conditional on their being infectious for *i* − 1 days. The grey arrows indicate the daily transition of individuals from one square to another that occurs with probability 1.

Our model is parameterised by *θ*, the probability of hospitalisation, Δ_*s*_, the daily probability of infection with strain *s*, and a discrete random variable *H* that characterises a set of probabilities governing daily transitions to hospital. Δ_*s*_ is informed by a discrete random variable, *Z*, that characterises a set of probabilities governing daily transitions into infected compartments, and is defined in Section 2.1.1. *Z* denotes the time in days from exposure of the infector to exposure of the infectee for a randomly chosen infectee-infector pair (i.e., generation interval), and can be viewed as the average relative contribution of each day to the individual reproduction number. The probability that infection occurs at infector age *i, ζ*_*i*_ = *P* (*Z* = *i*) = *F*_*Z*_(*i*) − *F*_*Z*_(*i* − 1), where the cumulative distribution function *F*_*Z*_(*i*) = *P* (*Z* ≤ *i*).

The event transmission of infection does not affect the individual’s stay in the compartment; however, for transition out of the *Y* compartments, the event going to hospital at infection age *i* is conditional on still being in the compartment at infection age *i* − 1. Given *H*, the random variable representing time from infection to hospital admission in hospitalised patients, we denote by *η*_*i*_ the probability of hospitalisation at infection age *i* given the individual was still not hospitalised at infection age *i* − 1, that is 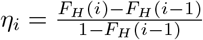. This is the discrete hazard of hospital admission at infection age *i* We use published estimates for *Z* and *H*, as described in section 2.1.3 below. Our model assumes that the proportion of infected people hospitalised and the processes governing hospitalisation and recovery over time are constant across strains.

#### 2.1.1 Infection dynamics

We extend a discrete epidemiological modelling framework by Sofonea et al. [7] to accommodate multiple viral strains spreading simultaneously. First, we express the effective density of infectious host population on a given day *d* that contributes to transmission of the strain *s* as:

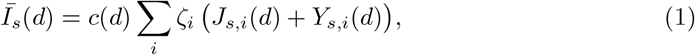

in which *J*_*s,i*_(*d*) + *Y*_*s,i*_(*d*) is the number of individuals with strain *s* on day *d* with infection age *i* in the community. Multiplying by *ζ*_*i*_ and summing over infection ages, _*i*_ *ζ*_*i*_(*J*_*s,i*_(*d*) + *Y*_*s,i*_(*d*)) can be regarded as the total ‘potential for infection’ in the community on day *d*. The effective infectious density, Ī_*s*_(*d*), is this sum scaled by the infectious contact ratio, *c*(*d*), on day *d*. The infectious contact ratio is the ratio of infectious contact rate on day *d* to infectious contact rate on day 0. The infectious density is thus a measure of the total amount of transmission in a completely susceptible population.

We allow susceptible hosts encounter the viral strain *s* with a probability Λ_*s*_(*d*), which as in [7] is assumed to follow the Michaelis-Menten function that saturates with the effective infectious density, *Ī*_*s*_(*d*) and the contact rate; under assumptions about initial conditions as in [7] we derive the following:

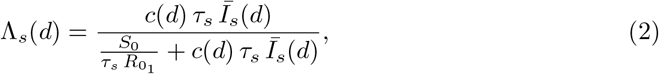

where *τ*_*s*_ is the relative transmission advantage of strain *s, S*_0_ the population size, and 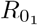 the basic reproductive number of the original strain. We define the *τ*_*s*_ as the ratio of the basic reproductive numbers of strain *s* to the original strain 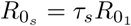.

When a host encounters multiple strains, we model the interaction between strains assuming superinfection with priority determined by order of exposure: i.e., only the first strain that encounters a host establishes infection when the same host subsequently encounters multiple strains. Thus, in the case of two strains, the probability of getting infected with the strain *s, s* ∈ {1, 2}, Δ_*s*_(*d*), is:

Δ_*s*_(*d*) = Pr(exposure to strain *s*) − Pr(exposure to both strains and *s*′ first)

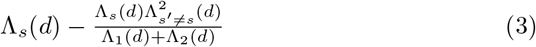

Similarly, expressions can be derived for more than two strains.

It follows then that the number of susceptibles on the next day is expressed as:

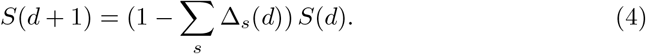

Of those exposed to either viral strain, the proportion *θ* will develop severe symptoms and eventually be admitted to the hospital (Fig. 1).

For less severe cases that do not result in hospitalisation, *J*, the infection progresses towards recovery until they are no longer infectious on the day *n*_*j*_:

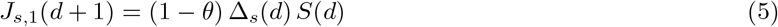

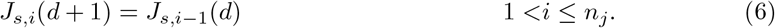

Those that develop severe symptoms, *Y*, are admitted to hospital with the probability *η*_*i*_ on the *i*-th day following exposure.

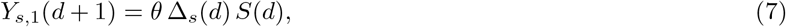

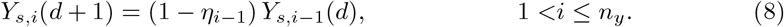

It follows then that the number of hospital admissions on day *d* + 1 equals

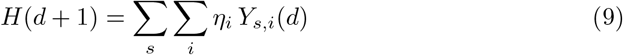

#### 2.1.2 Observed longitudinal data

Epidemiological models are often fitted to data on infected cases - however, case data depends on levels of testing, which varied over time during the COVID-19 pandemic. It can also be problematic to rely on data on deaths - for instance, many deaths in the ROI occurred following outbreaks in care homes, and thus data on deaths may not reflect disease spread in the general community. Biases and uncertainty in estimating the reproductive number arising from such issues are discussed elsewhere [6]. Thus, hospital admission data is likely a better reflection of the community spread of SARS-CoV-2. We used daily COVID-19 hospital admissions in the ROI and NI, reported respectively by the Central Statistics Office COVID data hub for the ROI [24], and the NI Department of Health [25].

Successive invasions of new variants have so far characterised the COVID-19 pandemic. Our study tracks two strains that circulated in the island of Ireland in the first 12 months of the pandemic: i.e., the original strain (initially detected in Wuhan, China) and the Alpha strain (also known as B.1.1.7., initially detected in Kent, UK). We used publicly accessible data on the frequency of the Alpha strain in the ROI [26], and NI [27], respectively.

#### 2.1.3 Incorporating empirical estimates of waiting time distributions

Linking transitions within and between these components are two random variables, each describing a waiting time process. These are the infectious period (generation interval), *Z*, and the delay between infection exposure and hospitalisation, *H*. The probability distributions representing these random variables have been estimated elsewhere empirically for SARS-CoV-2 in a global and European context.

##### Generation interval

The generation interval refers to the time between infection events in a pair of infector and infectee, reflecting the incubation duration and recovery timing. Here, we used the distribution of this interval to model the relationship between the age of infection (i.e., time since exposure) and the infectiousness of the infector. We employed an estimate by Ferretti et al. [28] who found the variation in SARS-CoV-2 generation interval was best described by the Weibull distribution with the mean interval of 5.5 days (shape= 3.29 and scale= 6.12). We truncated the Weibull distribution at the upper-integer-rounded 99%-quantile — without this truncation, the discrete model would require infinite time-tracking sub-compartments due to a right-unbounded support [0, ∞]. We then discretised the distributions because the dynamics unfold in discrete-time intervals of one day in our model.

##### Exposure to hospital admission

The waiting time between exposure and hospital admission was estimated as the sum of the incubation period and the delay between symptom onset and hospitalisation. We assumed that the two waiting times were independent due to the absence of evidence otherwise. A meta-analysis of global, but predominately, Chinese data found that the SARS-CoV-2 incubation period was log-normally distributed with parameters *µ* = 1.63 and *σ* = 0.50 [29], corresponding to a mean incubation time of 5.78 days (standard deviation of 3.97 days). The distribution of waiting time between symptom onset and hospitalisation was estimated assuming a gamma distribution by Public Health England with a mean of 5.14 days (standard deviation of 4.2 days) [30]. We fitted a gamma distribution to the simulated sum of the two distributions to represent the timing between exposure to infection and hospital admission (shape = 4.76 and rate = 0.435).

#### 2.1.4 Weekly infectious contact ratio

Here, we defined the contact ratio *c* as the (potentially infectious) contact rate relative to the pre-pandemic, pre-intervention baseline (eq. 1 & 2) and estimated this quantity using a piece-wise function consisting of weekly intervals. Specifically, we estimated the ratio in each area *a* (NI and ROI), per week *w* (i.e., *c*_*a,w*_) as a function of *ϕ*_*a,w*_, the log proportional change in the contact rate from the previous week. We index *w* from the date of the first public health intervention in either jurisdiction, which took place in ROI on 2020-03-12 (Supporting Information S1: Table S1 & S2); hence the preceding, pre-intervention contact ratios are defined as 1.0.

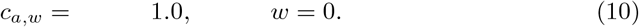

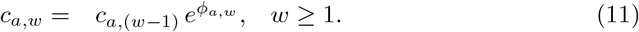

With this formulation, hierarchical Bayesian inference with priors on the *ϕ*_*a,w*_ allows us to estimate the time-varying weekly contact ratios with minimal prior information specific to the modelled system. Specifically, we used a prior ∼ 𝒩 (0, *ϵ*), where *ϵ* is a hyperparameter specifying the standard deviation of *ϕ*, such that *c*_*a,w*_ would equal *c*_*a*,(*w−*1)_ in the absence of signals from epidemiological data (Table 1). A priori, this formulation avoids over-fitting random weekly variation at the potential risk of smoothing over valid signals of an abrupt change in the weekly contact ratio, for example, following an introduction of lockdown measures. To check for such bias, we examined the extent to which our smoothing approach affects the estimation of sudden changes in the infectious contact ratio, *c*. We showed that our formulation is unlikely to introduce substantial bias (Supporting Information S2).

**Table 1:** Description of model parameters and their fixed values, or prior distributions used in Bayesian statistical inference. We assigned an informed prior for *R*_0_, *τ*_2_ and a generic, weakly informative prior for *Ī*_*s,a*_(0), ϵ and measurement error parameters.

| Symbol | Description | Fixed value or prior | Source |
| --- | --- | --- | --- |
| Epidemiological parameters |  |  |  |
| $\zeta$ | Infectivity | detailed in text | [36] |
| $\eta$ | Exposure to hospital admission | detailed in text | [29, 30] |
| $S_{0,ROI}$ | Population size of the Republic of Ireland | $4.92 \times 10^6$ | [37] |
| $S_{0,NI}$ | Population size of Northern Ireland | $1.89 \times 10^6$ | [38] |
| $\log(\bar{I}_{s,a}(0))$ | Log initial effective infectious density (of strain $s$ in area $a$) | $\mathcal{N}(0, 10)$ | |
| $R_{01}$ | Basic reproductive number of the original strain | $\mathcal{N}(2.79, 0.86)$ | [39] |
| $\tau_2$ | Transmission advantage of the Alpha strain | $\mathcal{N}(1.31, 0.24)$ | [32] |
| $\phi_{a,w}$ | Log proportional change in the contract ratio from the previous week | $\mathcal{N}(0, \epsilon)$ | |
| $\epsilon$ | Hyperprior specifying the standard deviation of $\phi$ | half-$\mathcal{N}(0, 1)$ | |
| $\theta$ | Probability of hospital admission given infection | 0.026 | [40] |
| Measurement error |  |  |  |
| $\sigma_h$ | Standard deviation for hospital admission (log-normal distribution) | half-$\mathcal{N}(0, 1)$ | |
| $\sigma_f$ | Standard deviation for frequency of the Alpha strain (beta proportion distribution) | half-$\mathcal{N}(0, 1)$ | |

#### 2.1.5 Initial conditions

The first case of SARS-CoV-2 on the island of Ireland was identified in NI on 2020-02-27 from an individual travelling back from Northern Italy via Dublin Airport located in ROI (Table S2). Two days later, the first official case in the ROI was also confirmed from a traveller from Northern Italy (Table S1). Initially, most known cases are travel-related, and contact tracing may successfully contain infections. As our model solely tracks community transmission, we started our simulations on the first day that community transmission was detected on the island of Ireland: 2020-03-05 (Table S1). Coincidentally, the exponential growth of confirmed cases appears to have begun around 2020-03-05 in both ROI and NI [24, HYPERLINK \l “bookmark33” 31]. We account for the uncertainty of the beginning of community transmission by estimating the initial infectious density independently in the two jurisdictions (Table 1). We assume implicitly that the contribution of the travel-related cases is negligible once the infection starts growing exponentially in the community.

The first cases of the Alpha strain were reported in November and December 2020, respectively, in ROI and NI (Tables S1 & S2). Due to high connectivity with the island of Britain, the Alpha strain likely entered the island of Ireland soon after it emerged in England, where the strain was detected in mid-September [32]. By February 2021, the Alpha strain comprised the majority of infections in both ROI and NI. To estimate the date of introduction, we fitted a three-parameter logistic function to the longitudinal data of the Alpha frequency and identified the date on which Alpha cases (frequency of Alpha × known new cases) intersects 1: the date of introduction was estimated as 2020-09-22. Again, we account for the sensitivity of the timing of introduction by estimating the founding infectious density of the Alpha strain, independently in the two areas (Table 1). In our model, viral strains differ only in their transmissibility, *τ*_*s*_.

#### 2.1.6 Fitting

We used a Bayesian approach to fit the above model to two types of longitudinal data from the ROI and NI: daily counts of hospital admissions and the Alpha strain frequency. Model parameters are detailed in Table 1. Hospital admissions per day were modelled as log-normally distributed with standard deviation parameters *σ*_*h*_. The frequency of the Alpha strain was fitted assuming the beta proportion distribution with a standard deviation parameter, *σ*_*f*_.

We fitted our model to the data from the first year of the pandemic from the first confirmed case of community transmission on the island, which was detected on 2020-03-05, in ROI, to the end of February 2021. Our modelling period precedes the widespread administration of the full course of vaccination in either jurisdiction: the proportion of fully (twice) vaccinated individuals in ROI and NI was less than 3% and 2% at the end of February 2021, respectively [24, HYPERLINK \l “bookmark33” 31].

Our model was written in Stan 2.21.2 and fitted through the RStan interface [33]. We fitted the model in parallel in four independent chains, each with 5000 sampled iterations and 1000 warmup iterations. For diagnostics, we confirmed over 400 effective samples and ensured convergence of independent chains using the 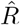 metric (values below 1.1 are considered an indication of multi-chain convergence) for all parameters [34]. We assessed the goodness of fit to data using standardised residuals (Supporting Information S3). We also quantified the posterior z-score and posterior contraction to examine the accuracy and precision of posterior distributions and the relative strength of data to prior information [35] (Supporting Information S4).

### 2.2 Counterfactual analyses

Estimating infectious contacts with a multi-strain model separates the effect of human behaviour from the difference in transmissibility of multiple strains. This separation allows us to leverage the epidemiological model and estimated parameters to simulate an epidemic based on data-generating processes consistent with the observed data. In turn, we can modify one part of the fitted model —while everything else is constant — to conduct counterfactual analyses, which allows us to explore the impact of different factors that affect disease transmission. Here, we explored two counterfactual scenarios: to examine the effect of lockdown timing; and to isolate the impact of the more transmissible Alpha strain on the hospitalisation outcome.

#### 2.2.1 Effect of lockdown timing

We explored the impact of the timing of lockdown introduction by simulating an epidemic with parameters estimated from the model fitted to the observed data on hospitalisations and strain proportions, but the contact ratios counterfactually shifted earlier by seven and 14 days relative to the actual start of the three lockdowns imposed in the ROI and NI. We then compared counterfactual scenarios and reality by computing the ratio of the cumulative hospital admission numbers for the subsequent days under the counterfactual versus the observed scenarios. Our effect measure is the (natural) log of this quantity, denoted lnRR.

Suppose the intervention is to shift the first lockdown date earlier by seven days. Denoting the actual lockdown date *d*_*l*_ and the counterfactual contact ratio on day *d* as *c*^*^(*d*):

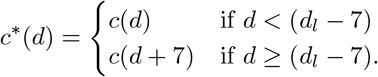

The counterfactual infectious density on day *d* follows from equation 1, and we denote this

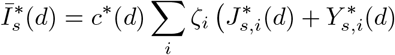

where the *J* ^*^, *Y* ^*^ denote the counterfactual numbers in these compartments on day *d*. From this follows the counterfactuals on day 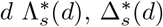, and *H*^*^(*d* + 1) from equations 2 to 9. For the second of third lockdowns, we assume that the epidemic had proceeded as observed up to the second and third lockdown, respectively.

#### 2.2.2 Impact of a more transmissible variant

We investigated the extent to which the introduction of the more transmissible Alpha strain contributed to public health burden by simulating an alternative epidemic with parameters estimated from the model fitted to the observed data on hospitalisations and strain proportions, but without introducing the Alpha strain in September 2020. We then used lnRR to compare the cumulative hospital admission numbers between the counterfactual and real scenarios over time until the end of the modelled period at the end of February 2021.

## 3 Results

### 3.1 Epidemiological model fit

Our discrete-time epidemiological model of SARS-CoV2 accurately described the timecourse of hospital admissions and the frequency of the Alpha strain during the first year of the pandemic in the two jurisdictions on the island of Ireland, before the full course of vaccines were widely administered (Fig. 2; Supporting Information S3 & S4).

**Figure 2:**
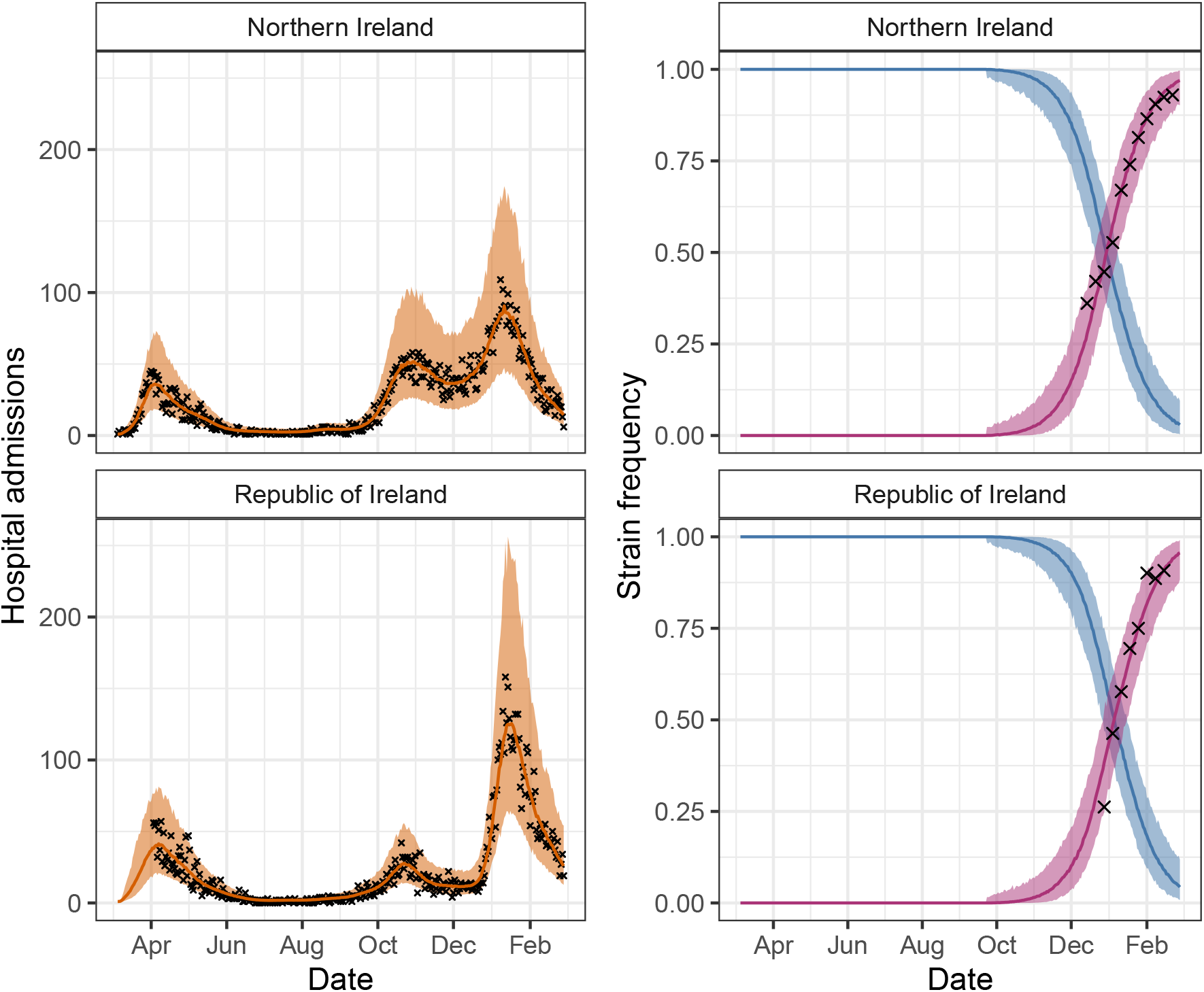
The fit of the epidemiological model to the longitudinal hospital admissions data and the frequency of the Alpha strain (the original strain in blue and Alpha strain in purple). The crosses indicate data and coloured bands correspond to 95% predictive intervals of the model, incorporating uncertainty in parameter estimation and sampling.

### 3.2 Estimated infectious contact ratios

We estimated a rapid decline in infectious contact ratios during the first month of the pandemic before a strict lockdown was implemented (Fig. 3; Tables S1 & S2). The first lockdown started on 2020-03-28 in both jurisdictions. By this date, the estimated infectious contact ratio was already down to about 60% of the pre-pandemic baseline in both jurisdictions. During this lockdown, estimated infectious contact fluctuated only slightly.

**Figure 3:**
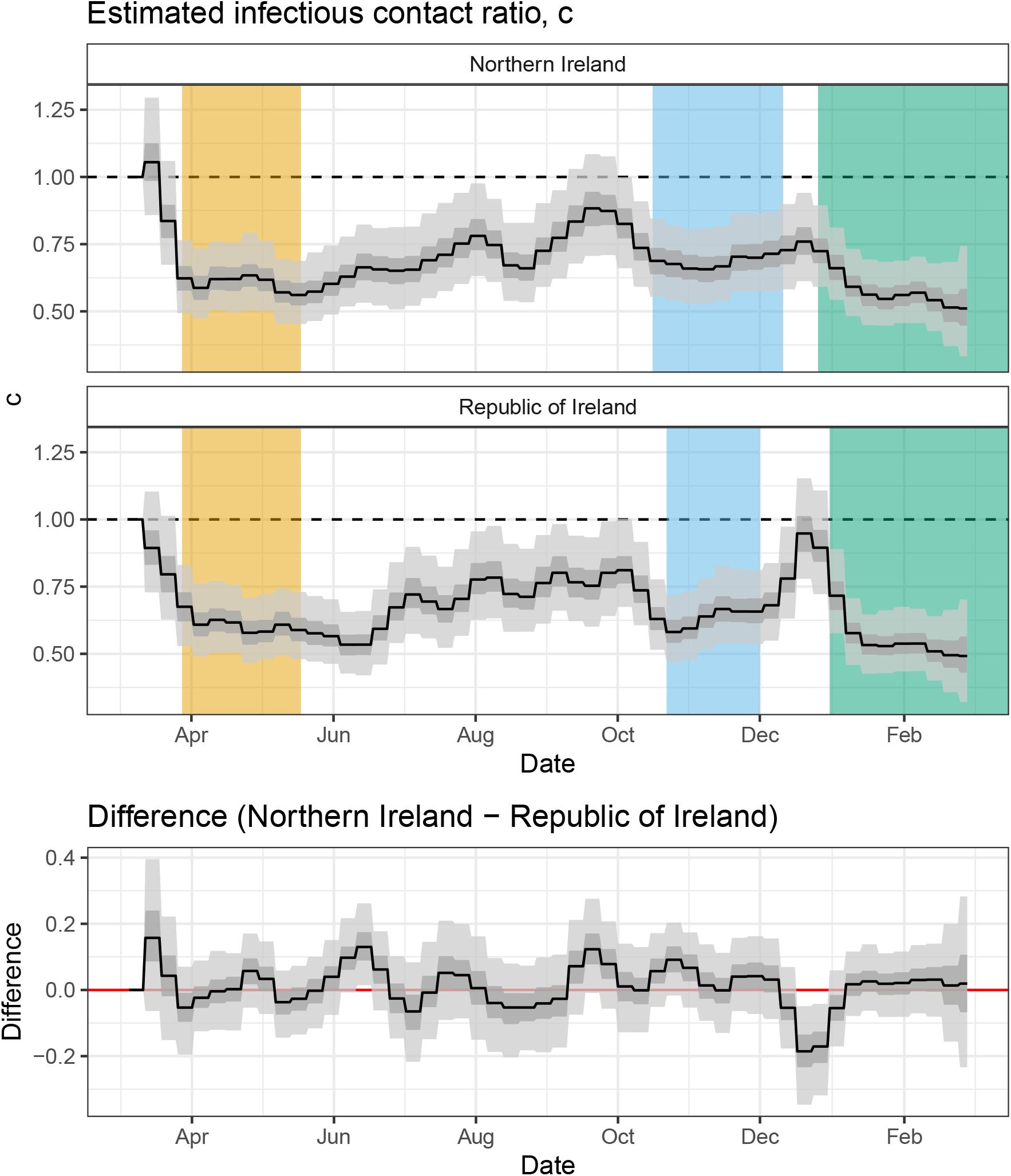
Estimated weekly infectious contact ratios in Northern Ireland (top) and the Republic of Ireland (middle) and differences in the contact ratio between the two jurisdictions (bottom). The three lockdown periods, corresponding to the most strict restrictions in each jurisdiction, are marked in yellow, blue and green, respectively. The black line, and grey bands correspond to the median, the 50% (dark) and 95% (light) predictive intervals.

In both jurisdictions, the easing of the first lockdown began from 2020-05-18, and a long period of slow restriction easing took place during the summer months. In the ROI, the estimated infectious contact ratio increased from June and fluctuated between approximately 70-80% of the pre-pandemic baseline in July, August and September. In NI, the contact ratio rose to a peak around the end of July. We detected higher infectious contact in NI than the ROI in mid-June (indicated by 95% predictive intervals of the difference excluding zero; Fig. 3; bottom panel). Of potential relevance, we note that all non-essential retail outlets were allowed to reopen earlier in NI than the ROI during this period from 2020-06-12 and 2020-06-29, respectively (Tables S1 and S2). Infectious contact in NI decreased through August but elevated again to about 90% of baseline by the end of September with no parallel increase in the ROI. This period corresponds to the first time primary and secondary teaching resumed in person in both jurisdictions. The increased contact in NI mirrors a trend in detected England where the September schooling reopening led to increased cases, most notably among the teaching staff [41].

Ahead of the second lockdown, the estimated infectious contact ratio declined to about 60% of baseline in both jurisdictions during October. Unlike during the other two lockdowns, the contact ratios tended to increase during the lockdown period in both jurisdictions throughout November (Fig. 3; top and middle panels).

At the beginning of December in the ROI, several mitigation measures were lifted, allowing non-essential businesses, restaurants, cafes and gastro-pubs to open as well as relaxing household gathering restrictions (Table S1). This period coincides with an increasing trend in the estimated infectious contact ratio, which reached about 90% of the pre-pandemic baseline the week before Christmas. In NI, on the other hand, the lockdown remained in place almost two weeks longer, and the estimated infectious contact ratio reached a maximum of about 75% of the baseline value before Christmas. Our estimates indicate that infectious contact was substantially higher in the ROI than NI for two weeks over the Christmas period (indicated by 95% predictive intervals of the difference excluding zero; Fig. 3; bottom panel): the ROI experienced the highest per capita infection rate in the world during this period. [42]. In both NI and the ROI, the third lockdown introduced in late-December 2020 coincided with the lowest contact ratio (Fig. 3; green) followed by the first lockdown in late March (Fig. 3; yellow) and second lockdown in November (Fig. 3; green).

### 3.3 Counterfactual scenarios

#### 3.3.1 Effect of lockdown timing

Lockdown measures have been shown effective in reducing the infection burden of SARS-CoV-2, and the timing of introduction is the most significant factor in determining their effectiveness [43, HYPERLINK \l “bookmark41” 44]. We found that bringing forward the lockdown dates by either seven or 14 days would have substantially reduced the cumulative hospitalisation over the subsequent 50 days from the date of lockdown in most scenarios (indicated by the 95% predictive interval of the lnRR excluding zero; Fig. 4). Of note, we found that a counterfactual simulation to bring forward the second lockdown date by seven days showed a non-conclusive impact on the cumulative hospitalisation in the subsequent 50-day period in either jurisdiction (judged by the 95% predictive interval of the lnRR containing zero; Fig. 4). The second lockdown was preceded by a declining trend in contact ratios while the contact during the lockdown remained relatively higher than the first or third lockdown (Fig. 3).

**Figure 4:**
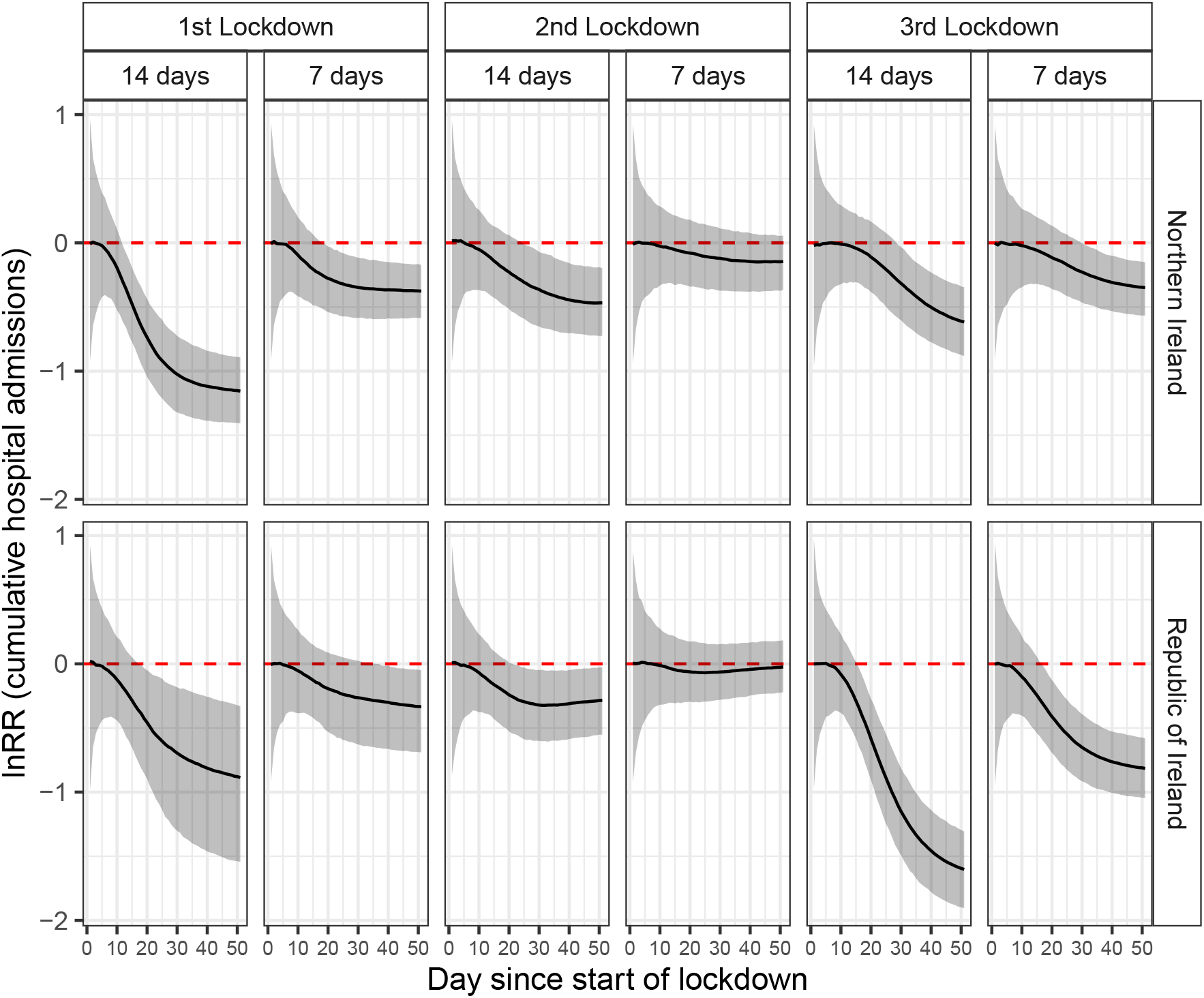
Counterfactual analysis demonstrates the effect of lockdown timing on epidemiological outcomes. The relative difference in cumulative hospital admissions between the counter-factual and factual scenarios is estimated as the log response ratio (lnRR). Shown are the lnRR following the introduction of three lockdowns in Northern Ireland and the Republic of Ireland, assuming that they would have started seven days and 14 days earlier. The black line and grey band indicates the median and 95% predictive interval, respectively.

#### 3.3.2 Impact of a more transmissible variant

Our model estimated that the Alpha strain was approximately 19% more transmissible than the original strain (95% prediction intervals [16.0, 21.8]). It is worthwhile noting that our model does not consider continuous inputs of infection into the island of Ireland, despite the high connectivity among the British Isles. Thus, our estimate of the Alpha transmissibility may be confounded by repeated introductions, for example, from England, where the Alpha strain was first detected. Nonetheless, our estimate is consistent with those from England [32].

To assess the the impact of the Alpha strain, which arrived later and is more transmissible than the original strain, we compared the fitted model (Fig. 3; orange) to a counterfactual simulation without the Alpha strain, in which we assumed the same estimated contact ratio (Fig. 3; blue). We detected a statistically distinguishable impact of the Alpha strain on the cumulative hospital admissions by earlier January in both jurisdictions - approximately 3.5 months after the initial introduction (indicated by the 95% predictive interval of lnRR excluding zero; Fig. 5). By the end of February 2021, we show that the Alpha strain was responsible for a 38 and 55% increase in cumulative hospitalisation, in NI (median lnRR = 0.323) and the ROI (median lnRR = 0.437), respectively (Fig. 5). Our findings demonstrate that an introduction of a more transmissible variant - without necessarily being more severe - can cause a large impact on the health care burden.

**Figure 5:**
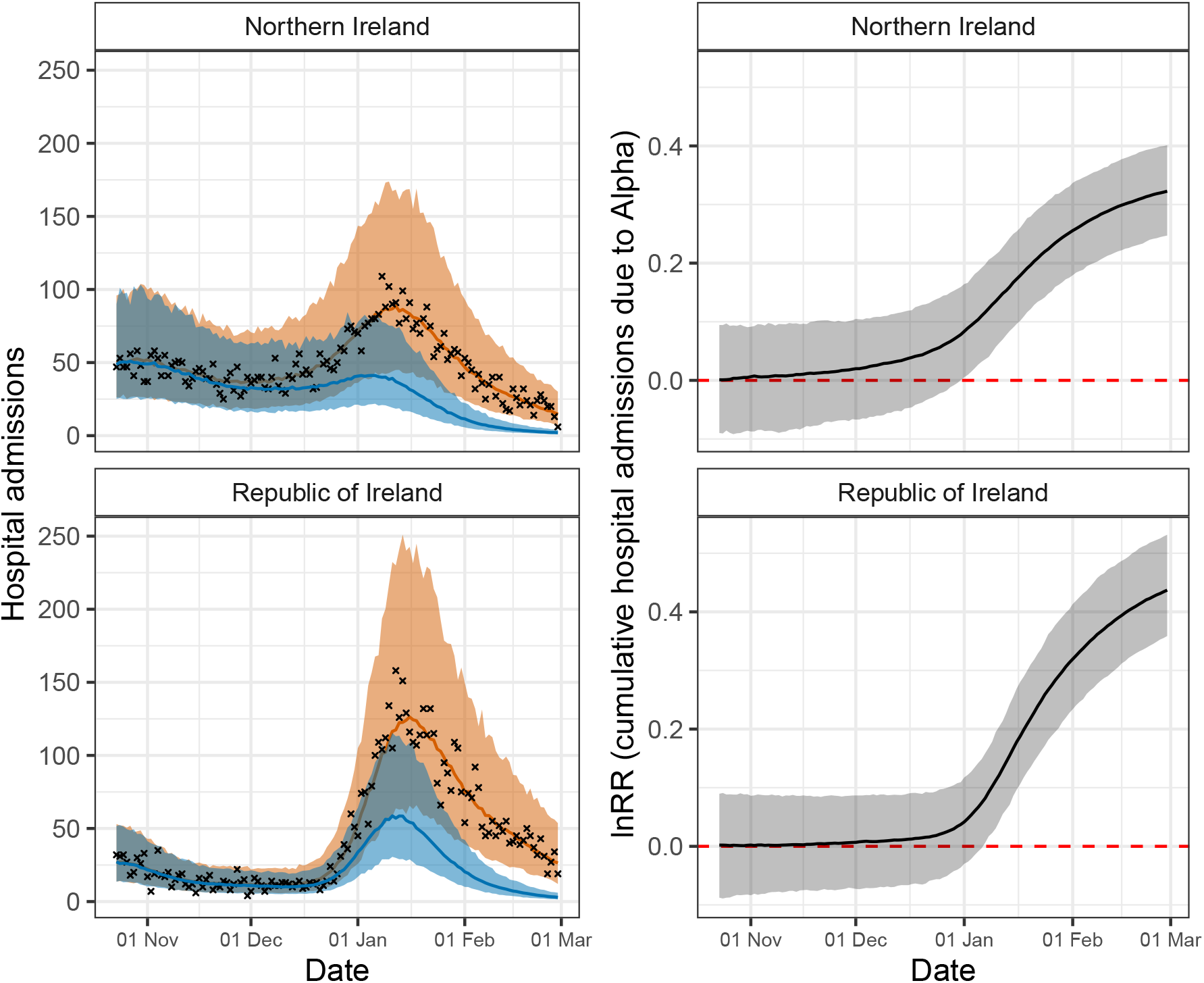
Counterfactual analysis shows the extent to which the Alpha strain elevated the burden of hospitalisation. To compute the impact of the Alpha strain, the counterfactual simulation (blue) assumes that the Alpha strain never invaded either jurisdiction. The crosses indicate data, and the coloured bands correspond to 95% predictive intervals of the fitted model (orange) and counterfactual scenario (blue), respectively, incorporating uncertainty in parameter estimation and sampling (left panels). The relative difference in cumulative hospital admissions between the two scenarios is estimated as the log response ratio (lnRR). The black line and grey band indicates the median and 95% predictive interval, respectively (right panels).

## 4 Conclusion

We developed a multi-strain model of SARS-CoV-2 and estimated time-dependent infectious contact over the first 12 months of the pandemic on the island of Ireland. Unlike many earlier COVID-19 modelling studies that estimate the effective reproductive number of a single strain, our model explicitly incorporates multiple viral strains and focus on estimating infectious contact ratios. An important difference between the infectious contact ratio and the effective reproductive number is that the former is unaffected by changes in virus transmissibility, which is modelled independently. As such, our approach separates the effect of human behaviour from that of the difference in transmissibilities between multiple, co-circulating strains.

Examining the longitudinal patterns and geographical differences in the estimated infectious contact ratios allowed us to identify corresponding policies and events. In addition, we leveraged estimated parameters to conduct counterfactual analyses, in which we examined the role of lockdown timing and a novel variant on cumulative hospitalisation. In a companion paper, we extended the application of the estimated infectious contact ratios to causal inference [45]. Specifically, we used mobility and mask-wearing data to independently predict the infectious contact ratios estimated from our epidemiological model described in the current paper and subsequently compared observed hospitalisations with predicted hospitalisations under a counterfactual mask-wearing scenario.

We presented a generic, epidemic model parameterised for SARS-CoV-2 to fit longitudinal hospitalisation data, one of the most reliable and available data types [6]. Of most relevance to COVID-19 in 2022, our model lacks human age structure and vaccination: these omissions give rise to certain limitations. For example, hospitalisation risks increase with age while older individuals adjust their behaviour differently from young counterparts [46]. Thus ignoring the age structure may bias our estimate of infectious contact estimated from hospitalisation data. In addition, the lack of vaccination and associated immunity in our model restricted our scope to the first 12 months of the COVID-19 pandemic. Technically, our model can be extended modularly to relax these assumptions about age structure and vaccination. However, these extensions were outside the scope of this study due to challenges in parameterising these processes reliably. For instance, the output of age-structured models are highly sensitive to assumptions of age-specific contact patterns [47], which likely changed during the epidemic, yet empirical data for time-dependent contact matrices are scarcely available. Behavioural adjustment in response to the pandemic is further complicated by the interaction between age- and sex-specific effects [48]. Furthermore, it is difficult to track and parameterise the state of immunity generated by natural infections from multiple viral strains and multiple vaccine doses using compartmental models.

Finally, our work contributes to the growing COVID-19 modelling literature by providing a transparent Bayesian workflow for fitting a multi-strain epidemic model to longitudinal epidemiological data, which may be readily adapted to modelling SARS-CoV-2 in other jurisdictions and other infectious diseases.

## Data Availability

All data produced in the present work are referenced in the manuscript.

