## Supporting Information for "Estimating time-dependent infectious contact: a multi-strain epidemiological model of SARS-CoV-2 on the island of Ireland"

### S1: Key events and mandated non-pharmaceutical interventions

Table S1: Key events and mandated non-pharmaceutical interventions in the Republic of Ireland. The week column refers to the week since the first public health intervention started in either jurisdiction (i.e., 2020-03-12 in the Republic of Ireland)

| Date | Intervention/Event | Week | Source |
|---|---|---|---|
| 2020-02-27 | First confirmed COVID-19 case in Northern Ireland returning from Italy via Dublin. | 0 | [1] |
| 2020-02-29 | First confirmed COVID-19 case in Republic of Ireland. | 0 | [2] |
| 2020-03-05 | First confirmed community transmission of COVID-19. | 0 | [3] |
| 2020-03-12 | Close all schools, colleges, and childcare facilities. | 1 | [2] |
| 2020-03-16 | Close bars and public houses. Advise against house parties. | 1 | [4] |
| 2020-03-28 | Stay at home order. | 3 | [2] |
| 2020-05-18 | Phase one easing of COVID-19 restrictions. | 10 | [5] |
| 2020-06-08 | Phase two-plus easing of COVID-19 restrictions. Stay local. | 13 | [5] |
| 2020-06-29 | Phase three easing of COVID-19 restrictions. | 16 | [5] |
| 2020-07-07 | COVID tracker contact tracing app is released. | 17 | [6] |
| 2020-08-08 | Co. Kildare, Laois, and Offaly are put into a stricter lockdown. | 22 | [7] |
| 2020-08-10 | Compulsory to wear a mask in shops and other enclosed public spaces. | 22 | [8] |
| 2020-08-18 | Work from home where possible. | 23 | [9] |
| 2020-08-21 | Co. Laois and Offaly specific restrictions lifted. | 24 | [10] |
| 2020-08-31 | Schools reopen. | 25 | [11] |
| 2020-09-16 | Five level framework introduced. National level 2 with some exceptions for Co. Dublin. | 27 | [12] |
| 2020-09-19 | Co. Dublin move to level 3. | 28 | [13] |
| 2020-09-26 | Co. Donegal move to level 3. | 29 | [14] |
| 2020-10-07 | National Level 3 with enhanced enforcement. Indoor dining in pubs and restaurants ban. | 30 | [15] |

| | | | |
|---|---|---|---|
| 2020-10-16 | Co. Cavan, Donegal and Monaghan move to level 4. Nationwide ban on household visits except for essential reasons. | 32 | [16] |
| 2020-10-22 | National level 5 lockdown. | 33 | [17] |
| 2020-11-02 | Alpha strain first detected. | 34 | [18] |
| 2020-12-01 | Reopen non-essential business. Households not to mix outside those within their bubble. No unnecessary inter-county travel. Face coverings mandatory in crowded areas. | 38 | [18] |
| 2020-12-04 | Reopen restaurants, cafes, gastropubs, and hotel restaurants for indoor dining with restrictions. | 39 | [19] |
| 2020-12-18 | Households can mix with up to two other households. Travel outside of your county permitted. | 41 | [19] |
| 2020-12-22 | Schools close. | 41 | [20] |
| 2020-12-24 | National level 5 lockdown with exceptions. | 42 | [21] |
| 2020-12-27 | No new inter-county travel. | 42 | [22] |
| 2020-12-29 | First vaccine administered. | 42 | [23] |
| 2020-12-31 | Level 5 lockdown with no exceptions. | 43 | [24] |
| 2021-01-04 | First calendar week after holidays to return to work where possible. | 43 | [25] |
| 2021-01-09 | Construction closes except for social housing and projects near completion. Negative COVID test required for arrivals from UK. | 44 | [26,27] |
| 2021-01-26 | Mandatory quarantine for all people arriving without a negative test. | 46 | [28] |

Table S2: Key events and mandated non-pharmaceutical interventions in Northern Ireland. The week column refers to the week since the first public health intervention started in either jurisdiction (i.e., 2020-03-12 in the Republic of Ireland)

| **Date** | **Intervention/Event** | **Week** | **Source** |
|---|---|---|---|
| 2020-02-27 | First confirmed COVID-19 case in Northern Ireland returning from Italy via Dublin. | 0 | [1] |
| 2020-03-20 | Close bars, gyms, restaurants and many other social venues. | 2 | [29] |
| 2020-03-28 | Lockdown and work from home. | 3 | [30] |
| 2020-04-24 | Opening of cemeteries on a restricted basis. | 7 | [30] |
| 2020-05-18 | Allowance to travel to garden and recycling centres. Marriage ceremonies allowed for terminally ill. | 10 | [30] |
| 2020-05-19 | Allowance to travel to places of worship for individual prayer. Attend church services, live music or theatre performances. Certain outdoor activities such as tennis, golf and angling allowed. Meet in groups of up to six people outdoors. | 10 | [30] |
| 2020-06-08 | People shielding able to spend time outside with people from their own household. Marriage/civil partnership ceremonies can take place outdoors (max 10 people). Reopen outdoor sports facilities and outdoor non-food retailers. Opening of non-food retail outlets. | 13 | [30] |
| 2020-07-03 | Hotels, bars and restaurants reopen subject to restrictions. Reopen tourist attractions and museums. | 17 | [30] |
| 2020-07-06 | Reopen hairdressers, spas, massage and reflexology providers. Visits to hospitals and care homes allowed to continue. | 17 | [30] |
| 2020-07-10 | Indoor gyms, bingo halls, amusement arcades, outdoor playgrounds and cinemas reopen. | 18 | [30] |
| 2020-07-24 | Number of people allowed to gather in a residential setting increased from six to ten. Reopen community centres, swimming pools and funfairs. Spectators are able to attend outdoor competitive games. | 20 | [30] |
| 2020-08-01 | Those shielding are now able to leave their homes. | 21 | [30] |
| 2020-08-03 | Eat Out to Help Out scheme was implemented. | 21 | [30] |
| 2020-08-10 | Compulsory to wear a mask in shops and other enclosed public spaces. | 22 | [30] |
| 2020-08-21 | People able to meet outdoors (max 15 people). Groups indoors limited to six people from two households. | 24 | [30] |

| | | | |
|---|---|---|---|
| 2020-09-14 | Local restrictions put in place in Belfast, Ballymena and parts of Glenavy, Lisburn and Crumlin. | 27 | [30] |
| 2020-09-22 | No longer able to visit others in their homes. | 28 | [30] |
| 2020-10-01 | Bars and restaurants close. | 30 | [30] |
| 2020-10-16 | Circuit breaker lockdown introduced. | 32 | [31] |
| 2020-10-19 | Schools to close for 2 weeks. | 32 | [30] |
| 2020-11-20 | Close contact services and unlicensed premises such as cafes able to open for 1 week. | 37 | [30] |
| 2020-11-27 | Non-essential retail close for 2 weeks. Places of worship to close. | 38 | [30] |
| 2020-12-08 | First vaccine administered. | 39 | [32] |
| 2020-12-11 | Reopen restaurants, gyms, non-essential retail and places of worship. Wet-pubs remain closed. | 40 | [30] |
| 2020-12-23 | Alpha strain first detected. | 41 | [33] |
| 2020-12-26 | Lockdown restrictions put back in place. | 42 | [30] |

### S2: Sensitivity to smoothing of the contact parameter, $c$

Our study tracks the infectious contact ratio, $c$ per week, to investigate the change in human behaviour. Specifically, we estimated the log proportional change in the contact ratio from the previous week, $\phi$ (Eq. 10 & 11, main text) with a prior $\sim \mathcal{N}(0, \epsilon)$, where $\epsilon$ is a hyper-parameter controlling the extent of smoothing: i.e., the lower the $\epsilon$ hyper-parameter, the larger the influence of $c$ from the previous week.

To examine the effect of smoothing on the estimation of infectious contact ratio $c$, we conducted a series of sensitivity analyses. First, we simulated data assuming three scenarios in which the contact ratio remains at 1.0 for two weeks following an introduction of SARS-CoV-2, and the ratio drops, on the third week, by 25, 50 and 75% respectively (green lines in Fig. S2). We then fitted the model (as described in Section 2.1.6 to the simulated data to assess whether the posterior distributions of $c$ tracks the "true" change in the contact ratio, $c$. We found that 95% highest posterior density (HPD) overlaps with the "true" contact ratio at 25% and 50% reduction in weekly contact (Fig. S1). When the contact ratio declines by 75%, the estimated contact ratio was overestimated for one week following an abrupt change (Week 3 at 0.75 in Fig. S1). We conclude that our approach to smooth the weekly contact ratio is unlikely to substantially bias interpretation as changes in human mobility per week are typically within 25% and unlikely to exceed 75% (Fig. S2).

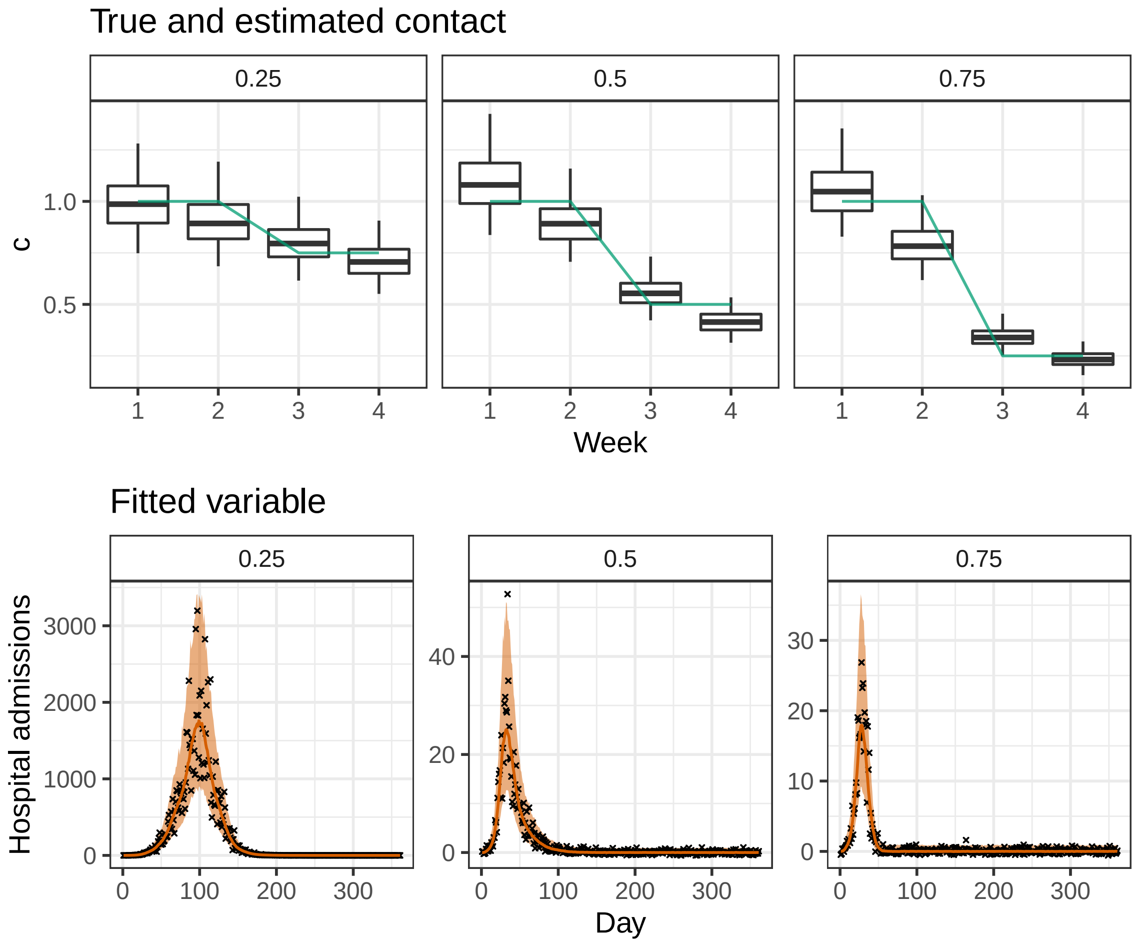


Figure S1: Smoothing of the contact parameter, $c$ based on Eq.10 & 11 (main text) is unlikely to bias inference of the 95% predictive intervals, so far as the weekly change in contact remains below 75%. We simulated data assuming three scenarios in which the contact ratio remains at 1.0 for two weeks following an introduction of SARS-CoV-2 and the ratio drops by 25, 50 and 75% respectively. The top panels show the "true" contact ratio (green line) and the estimated contact ratios (box and whiskers) over two weeks before and after the change in the contact ratio (i.e., between Week 2 and 3). With each box, the thick horizontal line, box edges and whiskers correspond to the median, 50% and 95% density of posterior distributions, respectively. The bottom panels show the simulated data (black crosses), and the coloured bands correspond to 95% predictive intervals of the model fit, incorporating uncertainty in parameter estimation and sampling.

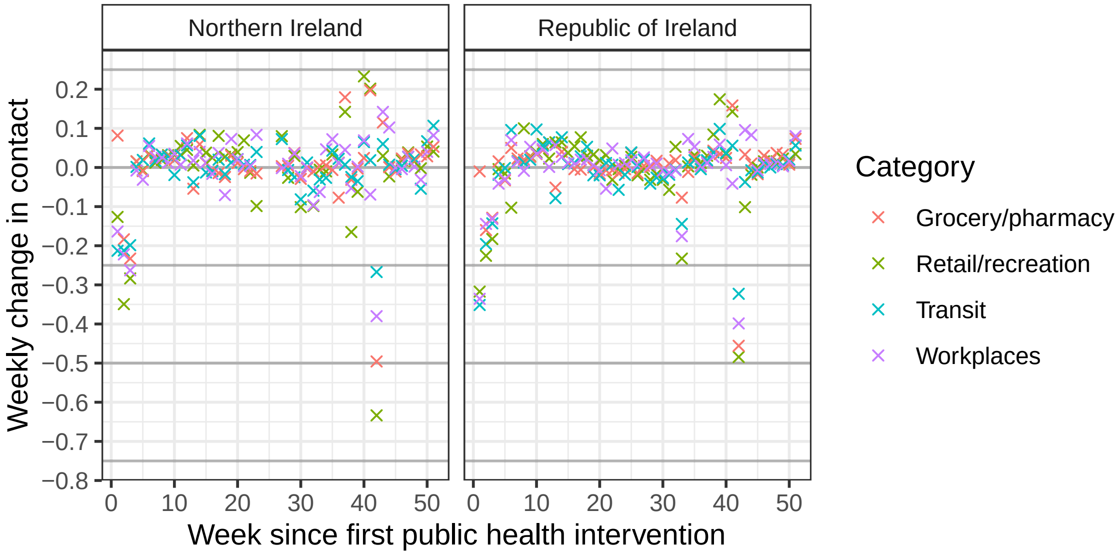


Figure S2: Google Mobility data indicate the weekly changes in contact are within 25% of the previous week in the majority of weeks. Shown are four categories of contact mostly likely related positively to SARS-CoV-2 transmission. The x-axis refers to the week since the first public health intervention was introduced on 2020-03-12 (Table 1 & 2).

#### S3: Assessment of model fit

To provide a rigorous assessment of the model fit, we examined the quantile of observed data with respect to the posterior prediction interval from the fitted model. We found the 95% posterior predictive interval frequently overestimates daily hospital admissions at very low numbers (i.e., less than or equal to 1; blue crosses in Fig. S3). For the rest, the 95% predictive interval contained over 95% of observed hospitalisation data. The Alpha strain frequency was accurately predicted by the 95% predictive interval in all observations.

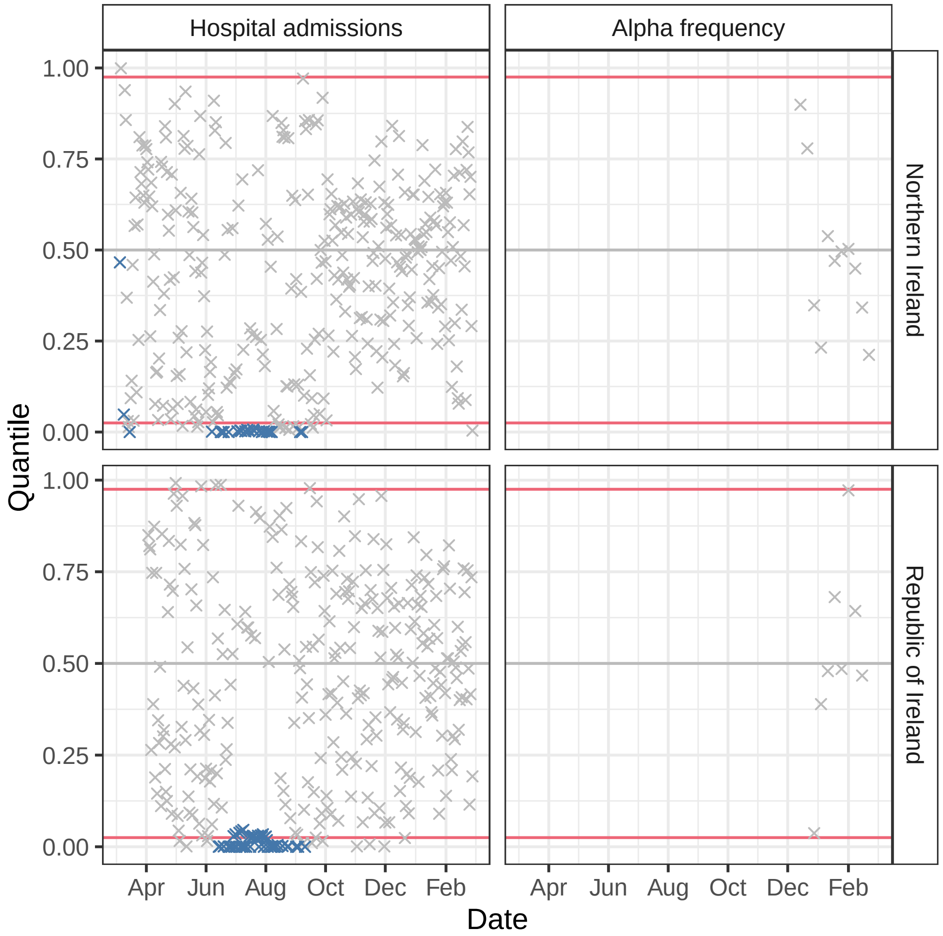


Figure S3: Quantile of observed data with respect to the posterior prediction interval from the fitted model. Poor fits are indicated by the observations (crosses) outside of the 95% posterior predictive interval (red lines). Daily hospital admissions less than or equal to 1 are indicated by blue crosses.

### S4: Assessment of posterior accuracy, precision and prior contraction

We leveraged the properties of posterior distributions to identify potential model fitting problems that might manifest from our model assumptions. To examine the accuracy and precision of posterior distributions, we first generated simulated observations based on the estimated posterior mean parameters. We then refitted our model to the simulated observations (i.e., secondary fitting) to compute the posterior z-score for each parameter, which measures how closely the posterior recovers the parameters of the data generating process [34]:

$$z = \frac{\mathbb{E}_{\text{sim}} - \mathbb{E}_{\text{post}}}{\sigma_{\text{sim}}},$$

where $\mathbb{E}_{\text{post}}$ denotes the posterior mean of the fit to the actual data that we consider the 'true' parameter. $\mathbb{E}_{\text{sim}}$ and $\sigma_{\text{sim}}$ denote the mean and standard deviation of the posterior distribution of the secondary fitting. The smaller the z-score, the closer the bulk of the posterior is to the true parameter [34]. In contrast, large z-values may be indicative of overfitting and, or poor bad prior specifications [34].

To examine the influence of the likelihood function in relation to prior information, we computed the posterior contraction, $k$:

$$k = 1 - \frac{\sigma^2_{\text{post}}}{\sigma^2_{\text{prior}}}$$

where $\sigma^2_{\text{post}}$ and $\sigma^2_{\text{prior}}$ correspond to the variance of posterior and prior distributions, respectively. The $k$ values close to zero indicate that data contain little information (i.e., rendering priors strongly informative). Conversely, values close to 1 indicate that data are much more informative than the prior [34].

We found that the most of our model parameters and hyperparameters — were estimated with accuracy and precision and identifiability, with the absolute posterior z-scores below three (Fig. S4). A small number of log proportional changes in contact, $\phi$ showed a tendency towards overfitting (the absolute posterior z-scores above three). The larger absolute posterior z-scores all coincided with periods of relative stasis in the contract ratio and included the weeks indexed 34 in NI and 6, 8, 9 and 49 for ROI (Fig. 3, main text; Tables S1 & S2). Thus, caution might be warranted when interpreting the contact ratio for these weeks — nonetheless, a small number of z-scores exceeding the absolute number of three is unlikely to be a cause of concern [34]. We found that the posterior distributions for $\phi$ contracted by over 50% compared to the prior distribution, i.e., $\sim \mathcal{N}(0, \epsilon)$, for most weeks, meaning that the weekly changes in contact were moderately informed by the smoothing hyper-prior $\epsilon$, which was itself well-identified from data (Fig. S4). The $\phi$ parameters were poorly identified for the last two weeks of the studied period (weeks indexed 50 and 51 in both justifications) as hospital admission data in the subsequent weeks required to inform these parameters were not included in model fitting.

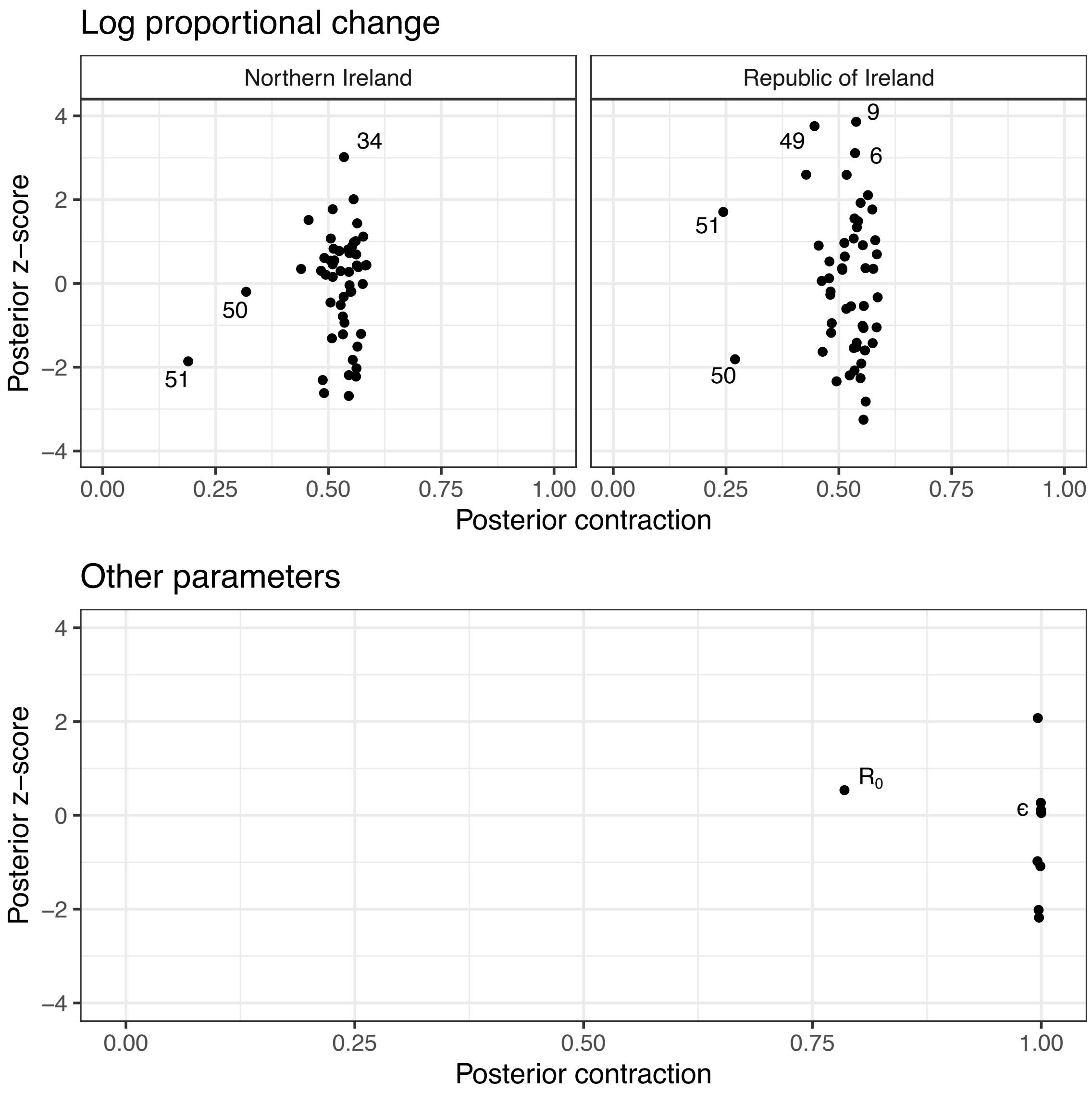


Figure S4: Accuracy, precision and identifiability of estimated parameters. Posterior z-score (y-axis) measures how closely the posterior recovers the parameters of the true data generating process and posterior contraction (x-axis) evaluates the influence of the likelihood function over the prior, respectively. Smaller absolute posterior z-scores indicate that the posterior accurately recovers the parameters of the data generating process: the absolute value beyond three to four may indicate substantial bias [34]. The posterior contraction values close to one indicate that data are much more informative than the prior. Other parameters refer to all estimated parameters excluding log-proportional changes (see Table 1 for the list of estimated parameters). The estimated parameter are represented by filled dots, which are labelled if notable.

[30] Department of Health Northern Ireland. COVID-19 mobility report;. Available from: https://www.health-ni.gov.uk/sites/default/files/publications/health/covid-19-mobility-reports-12-03-21.pdf.

[31] Executive Office Northern Ireland. Executive tightens restrictions to curb Covid-19; 2020. Available from: https://www.executiveoffice-ni.gov.uk/news/executive-tightens-restrictions-curb-covid-19.

[32] BBC News. Coronavirus vaccine: First care home residents in NI receive Covid jab; 2020. Available from: https://www.bbc.com/news/uk-northern-ireland-55216347.

[33] The Irish Times. First case of UK variant strain of Covid-19 confirmed in Northern Ireland; 2020. Available from: https://www.irishtimes.com/news/ireland/irish-news/first-case-of-uk-variant-strain-of-covid-19-confirmed-in-northern-ireland-1.4444730.

[34] Betancourt M. Towards a principled Bayesian workflow; 2020.
